# Evaluation of the impact of PEPFAR transition on retention in care in South Africa’s Western Cape

**DOI:** 10.1101/2023.01.20.23284819

**Authors:** Jessica Chiliza, Alana T Brennan, Richard Laing, Frank Goodrich Feeley

**Affiliations:** Department of Global Health, School of Public Health, Boston University, Massachusetts, Boston, USA; School of Public Health, University of Western Cape, Bellville, South Africa

**Author notes:** **(Corresponding author)** Jessica Chiliza, Address: 60 Stirling Crescent, Durban North 4051. **Disclaimer:** The author(s) of this article are solely responsible for the content.

**Keywords:** PEPFAR, HIV, retention, South Africa, low-and middle-income country

## Abstract

**Background:** Research on the impact of the PEPFAR transition in South Africa (SA) in 2012 found varying results in retention in care (RIC) of people living with HIV (PLWH).

**Objectives:** We investigated the factors that impacted RIC during the U.S. President’ s Emergency Plan for AIDS Relief (PEPFAR) transition in Western Cape, South Africa in 2012.

**Methods:** We used aggregate data from 61 facilities supported by four non-governmental organizations from to 2007-2015. The main outcome was RIC at 12-months after antiretroviral therapy initiation for two time periods. We used quantile regression to estimate the effect of the PEPFAR pull-out and other predictors on RIC. The models were adjusted for various covariates.

**Results:** Regression models (50^th^ quantile) for 12-month RIC showed a 4.6% (95%CI: -8.4, - 0.8%) decline in RIC post direct service. Facilities supported by Anova/Kheth’ impilo fared worst post PEFPAR; a decline in RIC of (−5.8%; 95% CI: -9.7, -1.8%), while that’ sit fared best (6.3% increase in RIC; 95% CI:2.5,10.1%). There was a decrease in RIC when comparing urban to rural areas (−5.8%; 95% CI: -10.1, -1.5%). City of Cape town combined with Western Cape Government Health facilities showed a substantial decrease (−9.1%; 95% CI: -12.3, -5.9%), while community health clinic (vs. primary health clinic) declined slightly (−4.4; 95% CI: -9.6, 0.9%) in RIC. We observed no RIC difference by facility size and a slight increase when two or more human resources transitioned from PEPFAR to the government.

**Conclusions:** When PEPFAR funding decreased in 2012, there was a decrease in RIC. To ensure the continuity of HIV care when a major funder withdraws sufficient and stable transition resources, investment in organizations that understand the local context, joint planning, and coordination are required.

## INTRODUCTION

Since 2003, President’ s Emergency Plan for AIDS Relief (PEPFAR) has been the United States’ most ambitious initiative to combat the global burden of HIV/AIDS and tuberculosis. PEPFAR is the largest contributor to global HIV/AIDS efforts (1). From 2004 to 2016, PEPFAR invested $72.7 billion globally in HIV and TB, including contributions to the Global Fund (2) PEPFAR has increased the number of people receiving HIV treatment globally,(3) and decreased HIV-related mortality by 10.5 % when compared to non-PEPFAR supported countries (4,5).

South Africa is the country with the highest number of people living with HIV (PLWH) globally (7.5 million).(6) By the end of 2017/2018 with 4.1 million adults on treatment (7), South Africa was running the largest HIV treatment program in the world (8). Due to the high burden of HIV/AIDS, South Africa was one of the first PEPFAR focus countries. In 2004, when PEPFAR began working in South Africa, the HIV prevalence among adults was 20% (9) and was a death sentence due to the lack of access to free care and treatment in public sector health facilities.

Initial PEPFAR funds were emergency funds spent on antiretroviral (ARV) treatment, using U.S. organizations based in South Africa (i.e., Population Services International, Family Health International) and private doctors (10) to roll out HIV treatment outside of the public health system (9). As time progressed during the direct service phase (2007-2012), PEPFAR supported local non-governmental organizations (NGOs) that employed health workers to work within the public health system to strengthen HIV services. In May 2009, the South African government (SAG)(11) adopted World Health Organization (WHO) treatment guidelines to start patients on treatment early (at a CD4 count of 350 rather than 200) (12), boosted the HIV budget by R1.7 million, and rolled out an ambitious testing campaign which reached 14.7 million South Africans in one year (12). During this time, PEPFAR strengthened its relationship with the SAG.

Most PEPFAR funds in South Africa were distributed to NGOs that work within state health facilities to strengthen HIV/AIDS care and treatment programs. In addition, many local NGOs were sub-contracted to various other organizations, resulting in PEPFAR partnering with 120 NGOs in South Africa by the end of 2013 (8).

### PEPFAR Transition in South Africa

SAG has been the main financial contributor to national HIV efforts. In 2012, to allow SAG to take greater financial responsibility for the South African HIV epidemic, the U.S government (USG) along with the SAG developed the Partnership Framework, which outlined an annual 48% funding decrease in PEPFAR funds ($483 to $250 million) by 2017 (13). The framework also outlined the transition of PEPFAR resources to the SAG and the USGs strategic shift from direct service (i.e., ARV roll out, purchasing ARV’ s and placing staff in SAG health facilities) to a focus on health systems strengthening, technical assistance, and sustaining health outcomes (14,15). In the Western Cape, human resource transition was a formalized process led by the Western Cape Government Health (WCGH) with input from health facilities. This resulted in the absorption of 78 HIV posts across the province (16).

There were multiple changes and challenges associated with this transition. Due to a change in PEPFAR leadership, the Partnership Framework did not develop as originally planned, PEPFAR’ s budget increased by 71%, from $259 million in 2015, to $443 million in 2016. The transition focused solely on care and treatment, and there was no plan for other PEPFAR-funded activities (i.e., prevention). Since the transition was negotiated at the national level, there was a lack of capacity at the provincial level to absorb PEPFAR-supported patients (17).

Literature on the impact of the PEPFAR transition in South Africa has found varying outcomes. Lince-Deroche et al. (18) looked at HIV service delivery post-PEPFAR in three clinics in Johannesburg and found no reduction in service delivery post-PEPFAR, while Cloete et al. (14) found 20% loss to follow up (LTFU) of patients transferred from private to government health facilities. Katz et al.’ s qualitative study found patients who were transferred to the public system were frustrated due to long queues and missed work opportunities, decreased quality of care, highlighting disrespectful staff, “low quality communication” and lack of holistic care (19). Kavanagh speculates approximately 50,000 to 200,000 people living with HIV were adversely affected by the PEPFAR transition (8). This high LTFU was a major concern due to lack of adherence and possible increase of drug resistant strains of the virus.

Retention in care (RIC) in South Africa (broadly defined as a patient’ s regular engagement with medical care at a health care facility after initial entry into the system) is a key indicator that demonstrates the long-term sustainability of ART programs. In 2015, the average RIC in Sub-Saharan Africa aligned with global RIC rates (74% RIC at 24 months).(20) However, in 2015, South African national RIC was slightly lower, at 57%) (21). More recent research using from South Africa’ s National Health Laboratory Service showed that HIV care retention was substantially higher (63.3%) when viewed from a national perspective than from a facility perspective (29%). Our results suggest that traditional clinical cohorts underestimate retention. Supporting the idea that failure to account for patient movement between clinics (sometimes referred to as a “silent transfer”) can make estimates of retention in care seem worse than they really are. To the best of our knowledge, there has been no formal evaluation of the PEPFAR transition in South Africa. As such, our study sought to assess the impact of PEPFAR transition on RIC in 2012.

## METHODS

### Data

The aggregate data used for this study was retrieved from the WCGH’ s HIV data system, (Tier.net) (22). The primary purpose of Tier.net is to manage the HIV program at a facility level. Data from Tier.net aggregated at the facility level were collected for 61 health facilities supported by four local PEPFAR treatment non-governmental organizations (NGO) from to 2007-2015: (1) Kheth’ impilo (n=15); (2) Anova Health Institute (Anova) (n=23); (3) Right to Care (n=5); and (4) TB, HIV/AIDS, Treatment Support, and Integrated Therapy (that’ sit) (n=11). There was a fifth category in our analysis of Anova/Kheth’ impilo, as these two NGOs overlapped in their support of seven health facilities in the study sample. Raw RIC data per health facility were provided for the cohort initiated ART each year. A cohort was defined by the WCGH as the number of new HIV patients (including transfers in) initiating ART treatment at a particular facility in a specific year (January 1-December 31) from 2007-2015.

### PEPFAR NGO’ s

The four NGOs used in this study were the main NGOs working in the Western Cape that received PEPFAR funding to support comprehensive HIV/AIDS care and treatment services at government facilities from 2007 to 2012 (Table 1). Note that Right to Care’ s timeline was slightly later, from 2009 to 2014. During this time, funding was used to scale up, support, and expand access to HIV services, including HIV testing and counselling, treatment, prevention of mother-to-child transmission, combination prevention and screening, and treatment of tuberculosis. In 2013, that’ sit, Right to Care and Kheth’ impilo received extension funds to close out projects and phase out direct service support starting in 2013 through 2015, while Anova and Kheth’ impilo received new PEPFAR grants to support the Western Cape from 2013 to 2017 in the Metro and Winelands regions. Each NGO worked in a specific geographic region in the province. As noted, Anova and Keth’ impilo both worked in the Metro District, supporting seven of the same health facilities.

**Table 1.** Western Cape NGO PEPFAR Timeline of Grants.

|  |  | Direct Service |  |  |  |  |  | Health Systems Strengthening |  |  |
| --- | --- | --- | --- | --- | --- | --- | --- | --- | --- | --- |
| NGO | District | 2007 | 2008 | 2009 | 2010 | 2011 | 2012 | 2013 | 2014 | 2015 |
| **Kheth'impilo** | Metro | x | x | x | x | x | x | Extension<br>+ New<br>Grant | x | x |
| **that'sit** | Eden | x | x | x | x | x | x | Extension | x | x |
| **Right to**<br>**Care** | Overberg; Central Karoo |  |  | x | x | x | x | x | Extension | x |
| **Anova** | West Coast | x | x | x | x | x | x |  |  |  |
| **Anova** | Winelands | x | x | x | x | x | x | New Grant | x | x |
| **Anova** | Metro | x | x | x | x | x | x | New Grant | x | x |

### Study variables

The main outcome of interest was RIC at 12 and 24 months after ART initiation in each health facility from 2007 to 2015. RIC was analyzed for two separate time periods: (1) PEPFAR direct service (2007 to 2012) and (2) post-PEPFAR direct service (2013 to 2015). The study definition for RIC among adults (age >15 years) is as follows:

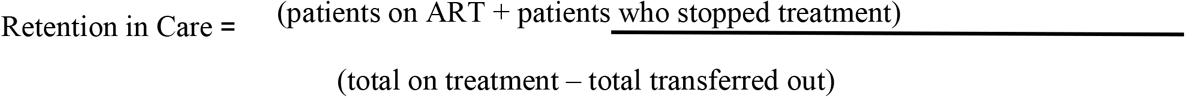

Total on treatment includes HIV clients who are transferred to the health facility via a formal or silent transfer. Mortality was included in patients who stopped treatment. We conducted a sensitivity analysis by adding death to the definition, and the results did not change. Retention was calculated across the nine-year period (2007-2015) for each clinic. Other covariates of interest were patient volume for each clinic (stratified into quintiles, location (urban vs. rural), number of job posts transferred from PEPFAR NGOs to government (categorized as ≤1 and ≥2), facility type (central day clinic (CDC), community health clinic (CHC), and primary health clinic (PHC)) and government (City of Cape Town, Western Cape Government and a combination where they both overlapped) and NGO (Anova, Anova/Kheth’ impilo, Kheth’ impilo, that’ sit and Right to Care). RIC for each health facility supported by Anova, that’ sit and Kheth’ impilo were calculated at 12 and 24 months for the two time periods 2007 to 2012 (during PEPFAR direct service) and 2013 to 2015 (post-PEPFAR direct service). Since the Right to Care became active two years later, the average RIC cut-off was 2009 to 2012 (PEPFAR direct service) and 2013 to 2015 (post-PEPFAR direct service).

Simple descriptive statistics were used to report the characteristics of the study sample and were stratified by NGO. We graphically displayed trends in 12- and 24-month retention, loss to follow-up (defined as clients who have not visited the health facility for more than 90 days), and total clients starting treatment at the start and end of the cohort by year overall and stratified by NGO.

Quantile regression was used to estimate the associations between PEPFAR pull-out and changes in 12-month retention in care at the 25^th^, 50^th^ and 75^th^ quantiles adjusted for covariates. The models contain the dependent variable (12-month retention), conditional on time (years), plus an indicator variable for PEPFAR pull-out (set to 0 for each year during PEPFAR funding and set to 1 for years post-PEPFAR), an interaction term between these two variables to display trends over time, and additional covariates (i.e., clinic volume, government, NGO, transfer in human resource posts, facility type, and location). The coefficient for the indicator variable for PEPFAR pullout can be interpreted as the quantile difference in retention between the PEPFAR and post-PEPFAR periods for the 25^th^, 50^th^ and 75^th^ quantiles of our study sample. We graphed the crude predicted results from the quantile regression models and overlaid scatter plots of retention by clinic overall and stratified by each NGO to show the intercept shift and trends in retention during and post-PEPFAR. We conducted quantile regression on 24-month retention as well but focused on the main results of this study on 12-month retention because they did not differ greatly and displayed 24-month retention as supplementary tables and figures. STATA 16 and Excel 2016 were used to analyze the data.

## RESULTS

### Characteristics of the study sample

The 61 health facilities included in the study sample were predominately WCGH owned (77%), with a total of 190,343 patients, and equally split between rural and urban areas (Table 2). The majority of the clinics were supported by Anova (n=23, 38%) and Kheth’ impilo (n=15, 25%), while Right to Care had the lowest number (n=5, 8%). ART cohorts gradually increased in size, though in 2015, there was a dramatic increase in the number of HIV clients on treatment (Figure 1).

**Table 2:** Study characteristics stratified by NGO (N=61)

|  |  | NGO<br>(n=total facilities supported) |  |  |  |  |  |
| --- | --- | --- | --- | --- | --- | --- | --- |
|  |  | Anova<br>n= 23 (37.7%) | Anova/Kheth'impilo<br>n=7 (11.5%) | that'sit<br>n=11<br>(18%) | Right to Care<br>n=5<br>(8.2%) | Kheth'impilo<br>n=15<br>(24.6%) | TOTAL |
| **Total facilities supported, n (%)** |  | 23 (38%) | 7 (11%) | 11 (18%) | 5 (8%) | 15 (25%) | 61 (100%) |
| **Total Clients on ART, n (%)** |  | 84,610 (44%) | 39,752 (21%) | 13,202 (7%) | 5,528 (3%) | 47,251 (25%) | 190,343 (100%) |
| **Government ownership of facilities, n (%)** | WCGH | 20 (87%) | 3 (43%) | 11 (100%) | 5 (100%) | 8 (53%) | 47 (77%) |
|  | CoCT | 2 (9%) | 3 (43%) | - | - | 7 (47%) | 12 (20%) |
|  | combined | 1 (4%) | 1 (14%) | - | - | - | 2 (3%) |
| **Geographic area, n (%)** | rural | 14 (61%) | - | 11 (100%) | 5 (100%) | - | 30 (49%) |
|  | urban | 9 (39%) | 7 (100%) | - | - | 15 (100%) | 31 (51%) |
| **12 month RIC PEPFAR Direct Service, % (95% CI)** |  | 65% (64.6-65.5%) | 67.8% (67.2%-68.5%) | 69.1% (68%-70.1%) | 75.8% (74.1%-77.4%) | 70.4% (69.9%-71%) | 67.5%(67.2-67.8%) |
| **12 month RIC Post PEPFAR Direct Service, % (95% CI)** |  | 57.7% (57.1-58.2%) | 60.8% (60.1%-61.6%) | 60.4% (59.1%-61.6%) | 66.4% (64.5%-68.2%) | 66.2% (65.5%-66.8%) | 61% (60.7%-61.4%) |
| **12 month Overall RIC (2007-2015), % (95% CI)** |  | 62% (61.6%-62.3%) | 64.7% (64.2%-65.2%) | 65.3% (64.4%-66.1%) | 71.1% (69.9%-72.4%) | 68.5% (68.1%-69%) | 64.7% (64.5-64.9%) |

**Figure 1.**
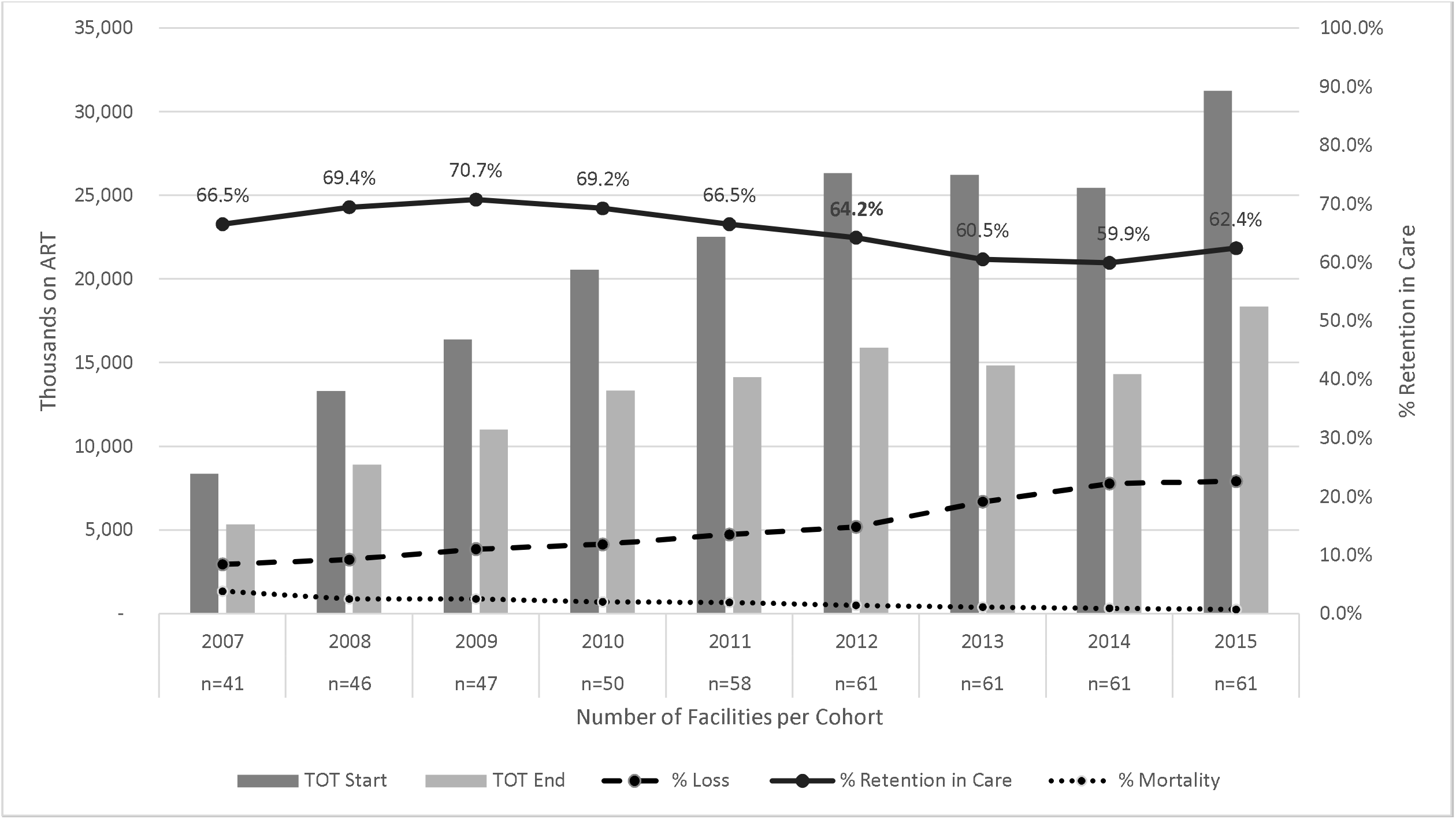
12-month retention in care, loss to follow-up and death overlaid on total patients on ART at the start and end of the 12-month period.

### Trends of retention over time

When assessing the trends in 12-month retention overall, our results show an increase in the number of people starting ART from 8,338 in 2007 to 31,260 in 2015 (Figure 1), with Anova having the greatest increase in patients during that time period and Right to Care having the smallest (Supplemental Figure 2a-2e). The overall 12-month retention of the study sample was 67.5% (95% confidence interval (CI): 67.2-67.8%) during PEPFAR and 60.8% (CI: 60.4-61.1%) post-PEPFAR. Retention for each NGO decreased post-PEPFAR direct service, with the exception of Anova, which observed an increase in RIC of 14.3% points post-PEPFAR. Overall, the graphs for each NGO showed a decrease in RIC in 2012/2013 (Supplemental Figure 2a-e).

**Figure 2.**
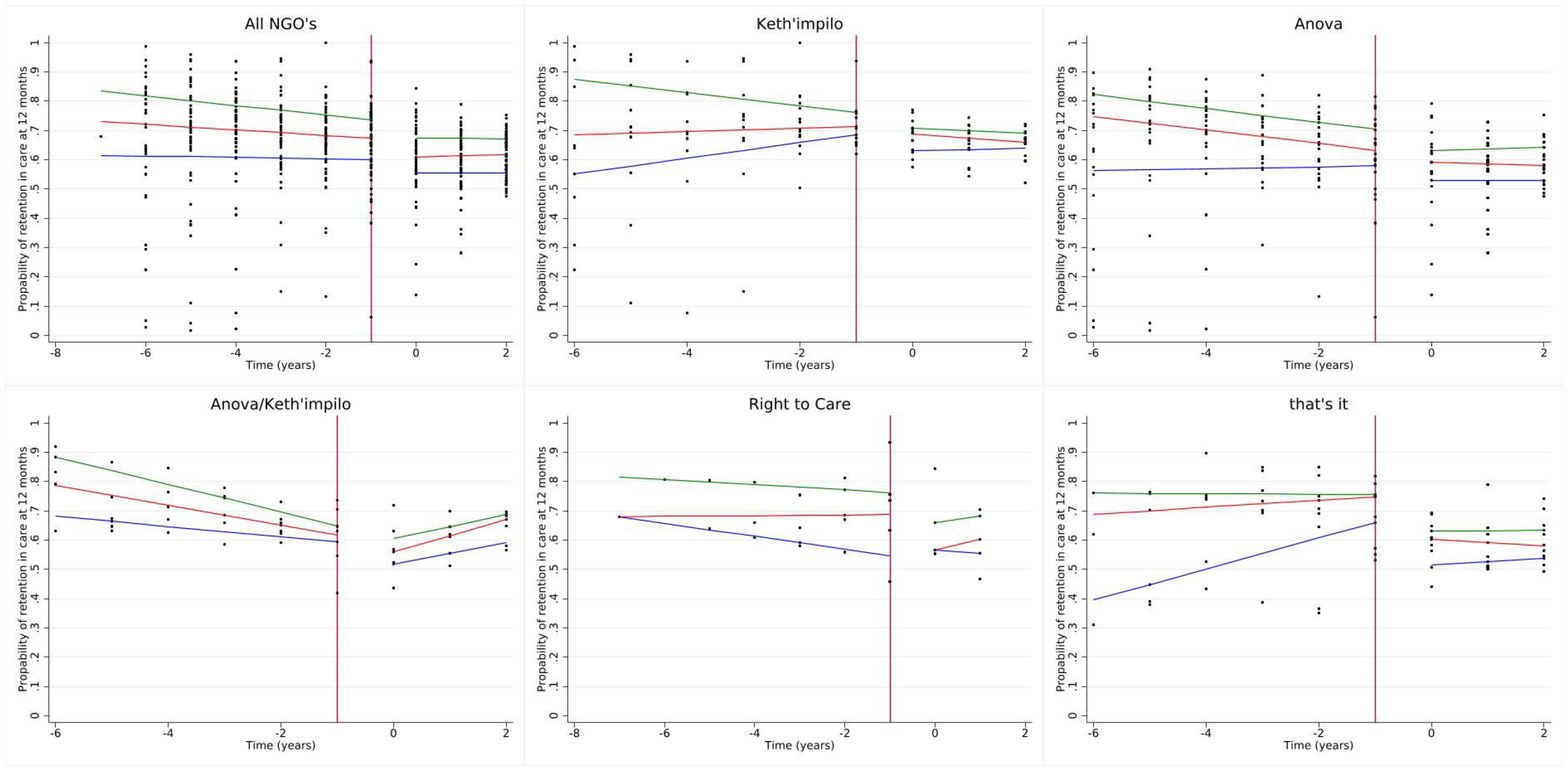
Quantile regression estimates for 12-month retention stratified by NGO.

The mortality rate of the study sample substantially decreased post the PEPFAR transition, falling from 3.8% in 2007 to 0.7% in 2015 (post-PEPFAR) (Figure 1). This is most likely due to a delay in reporting of mortality, as previous research shows 50% of loss to follow-up is due to mortality.(23)

### Quantile regression

We report the results for the 50^th^ quantile for all models, as the 25^th^ and 75^th^ percentiles are displayed in the tables and figures. The median RIC for the 50^th^ percentile was 66.0% (interquartile range (IQR): 57, 73%). We observed a decline of 4.0% (quantile difference (QD) - 7.7, -0.4%) in 12-month RIC post-PEPFAR compared to during PEPFAR (Table 3 and Figure 2). Additionally, there was a slight decline in RIC in the PEPFAR era and a plateau in the post-PEFAR period (indicated in the direction of the slopes), as shown in Table 3 and Figure 2. It is also important to note that, health facilities supported by Anova/Kheth’ impilo fared worst with regard to RIC post PEFPAR (50^th^ QD: -4.9%; 95% CI:-8.8, -1.0%), while that’ sit fared best (50^th^ QD: 3.6%; 95% CI: -0.2, 7.3%) when compared to Anova. We observed a larger decline post-PEPFAR in 24-month RIC (50^th^ QD: -7.0%; 95% CI: -10.2, -3.9%) and all other NGO’ s performing slightly better post-PEPFAR when compared to Anova (Supplemental Table 1 and Supplemental Figure 2a-f.)

**Table 3:** Crude and adjusted quantile regression for the outcome of 12-month retention.

|  | Crude quantile difference (95% CI) |  |  | Adjusted quantile difference (95% CI) |  |  |
| --- | --- | --- | --- | --- | --- | --- |
|  | 25% | 50% | 75% | 25% | 50% | 75% |
| **post PEPFAR** | -4.2% (-11.6, 3.2%) | -5.6% (-10.4, -0.7%) | -4.5% (-8.6, -0.4%) | -5.2% (-12.4, 2.0%) | -4.6% (-8.4, -0.8%) | -4.8% (-9.2, -0.3%) |
| **slope during PEPFAR** | -0.2% (-1.7, 1.2%) | -1.0% (-1.9, 0.0%) | -1.6% (-2.4, -0.9%) | 0.5% (-0.9, 2.0%) | -1.3% (-2.1, -0.6%) | -1.9% (-2.8, -1.0%) |
| **change in slope post-PEPFAR** | 0.0% (-2.7, 5.7%) | -0.5% (-3.3, 4.3%) | -0.2% (-3.3, 3.0%) | 0.0 (-5.7, 5.5%) | -0.3% (-3.3, 2.7%) | 2.5% (-0.1, 5.0%) |
| **Constant** | 59.8% (54.4, 65.2%) | 66.4% (62.9, 70.0%) | 72.0% (69.1, 75.0%) | 57.3% (47.3, 67.3%) | 73.2% (67.9, 78.5%) | 81.9% (75.7, 88.1%) |
| **PEPFAR NGO** |  |  |  |  |  |  |
| Anova | - | - | - | ref. | ref. | ref. |
| Kheth'impilo | - | - | - | 6.1% (-0.6, 12.8%) | -1.6% (-5.2, 2.0%) | -1.2% (-5.3, 3.0%) |
| Anova/ Kheth'impilo | - | - | - | 0.4% (-7.0, 7.9%) | -5.8% (-9.7, -1.8%) | -7.5% (-12.2, -2.9%) |
| Right to Care | - | - | - | 7.7% (-1.9, 17.4%) | 6.0% (0.8, 11.1%) | 6.6% (0.6, 12.6%) |
| that'sit | - | - | - | 1.9% (-5.2, 9.1%) | 6.3% (2.5, 10.1%) | 5.5% (1.1, 10.0%) |
| **clinic size** |  |  |  |  |  |  |
| 174-1020 | - | - | - | ref. | ref. | ref. |
| 1021-2562 | - | - | - | 6.6% (-0.6, 12.8%) | 4.9% (1.1, 8.7%) | -0.0% (-4.5, 4.4%) |
| 2563-4021 | - | - | - | 4.3% (-3.0, 11.7%) | 1.6% (-2.3, 5.5%) | -3.6% (-8.1, 1.0%) |
| 4022-5856 | - | - | - | 9.5% (2.0%, 16.9%) | 3.5% (-0.5, 7.4%) | -0.3% (-4.9, 4.4%) |
| 5857-9760 | - | - | - | 11.4% (2.5, 20.2%) | 2.1% (-2.6, 6.8%) | -5.6% (-11.1, -0.1%) |
| **location** |  |  |  |  |  |  |
| rural | - | - | - | ref. | ref. | ref. |
| urban | - | - | - | 5.0% (-3.1, 13.0%) | -5.8% (-10.1, -1.5%) | -8.9% (-13.9, 3.9%) |
| **government support** |  |  |  |  |  |  |
| City of Cape Town | - | - | - | ref. | ref. | ref. |
| Western Cape Government | - | - | - | 2.7% (-8.6, 13.9%) | -3.2% (-9.2, 2.7%) | -4.6% (-11.6, 2.4%) |
| Combined | - | - | - | -8.3% (-14.3, -2.2%) | -9.1% (-12.3, -5.9%) | -7.5% (-11.2, -3.8%) |

We also saw a large decrease of -7.8% (95%CI: -12.8, -2.9%) for the 50^th^ quantile in RIC in urban health facilities when compared to rural areas, while City of Cape Town (CoCT) combined with WCGH had a substantial drop (50^th^ QD: -6.1%; 95% CI:-10.6, -1.7%) in RIC when compared to CoCT alone, while CHCs (50^th^ QD: -6.4%; 95% CI: -10.6, -2.1%) and PHCs (50^th^ QD: -2.8%; 95% CI: -5.8, 0.1%) had a larger decline in RIC when compared to CDCs. We saw an increase of RIC in health facilities that had ≥2 posts transferred from PEPFAR to government (50^th^ QD: 3.4%; 95% CI: -0.1, 7.0%) compared to ≤1. There was no clear trend in RIC when assessing the relationship between health facility size, measured by total clients on ART, and RIC. With regard to the 24-month RIC, we saw a slight decline in RIC as health facility size increased, while trends in all other variables were consistent with the 12-month RIC (Supplemental Table 1 and Supplemental Figure 2a-e).

## DISCUSSION

In this assessment of RIC post-PEPFAR in South Africa’ s Western Cape, we found a 12-month retention decline in the post-PEPFAR era. For all NGO supported clinics, RIC during PEPFAR was a 6.7% percentage point decrease from the PEPFAR direct service period. There are several explanations for these results. One explanation for the decline from 2012 could be the direct effect of PEPFAR, moving from direct service support to a focus on health systems strengthening. This change meant a decrease in human resources, supported by the local healthcare system. Much of PEPFAR’ s support was for HIV-specific community health workers, tracers, and data capturers, who were key for high performance and sustained HIV outcomes, particularly retention. The Western Cape PEPFAR transition did not prioritize the absorption of PEPFAR-supported community posts, which meant that in 2012/2013, the province lost 418 community posts supported by PEPFAR (16).

Our findings are consistent with Kavanagh’ s (8) report on the South African transition. Owing to PEPFAR’ s transition strategy, health facilities lost close ties with the community, and HIV retention and prevention efforts fell off the priority list. If the government had made the decision to transition to more staff in smaller health facilities and prioritize community staff in the transition, they would have been more likely to see sustained retention in smaller health facilities. Another alternative explanation could be the 2010 change in HIV treatment policy, which increased the CD4 eligibility threshold for ART from 200 to 350 cells/mm^3^ (24) The increase in eligibility threshold would have resulted in more patients accessing the health care system for monitoring and treatment of the HIV disease, stressing the health system and beginning ART with more patients who were not yet seriously symptomatic. Research from Cape Town, South Africa showed that LTFU and the risk of virological failure increased when the ratio of patients per health worker increased (25). One explanation for this could be the increase in ARV treatment sites over time and that patients were able to move between facilities but would be recorded as LTFU in the original health facility where they were enrolled on treatment.

When stratifying our results by NGO, we showed that overall RIC at health facilities supported by Kheth’ impilo was the highest, while Anova supported health facilities with the lowest RIC (Table 2). Kheth’ impilo support was localized to urban areas, and even with high ART patient volumes, they were able to maintain high RIC throughout the transition. Kheth’ impilo was also able to retain the largest number of PEFPAR posts (28) compared to the other NGOs. Sustaining former PEPFAR human resources in the health system likely facilitated high RIC (26).

The CoCT RIC performance was better than that of WCGH. CoCT may have performed better than WCGH health facilities because post-Apartheid or post 1994 they inherited a relatively strong health system that provided HIV services for longer than the WCGH and spent more funding per patient than WCGH (27). Pre-1994 and prior to PEPFAR, HIV care and treatment were not available. HIV treatment was initially rolled out in the Metro District, leaving the rural areas of the Western Cape with little access to HIV services. Additionally, because CoCT provides services in an urban area, there is easier access to healthcare services, allowing patients to stay on their treatment. HIV patients living in rural areas often lack transport and resources and fear loss of confidentiality, while rural health systems suffer more medication stockouts, which affect RIC (28).

Anova/Kheth’ impilo were impacted more, with as high as a 7.5% decline in RIC post-PEPFAR in the 75^th^ quantile vs. Anova alone, while Right to Care was the least impacted, with as high as a 7.7% increase post-PEPFAR in the 75^th^ quantile vs. Anova alone. We recently published a qualitative analysis of this work (26) which showed that established NGOs with a history of working in Western Cape supported facilities with higher RIC. Anova and Kheth’ impilo had been working in Western Cape for many years, understood the health system gaps, had long-standing relationships with local officials, and produced high sustainability results. Qualitative data highlighted that stable staff and the consistency of patient/provider relationships were important for sustaining RIC. It is important that patients trust and feel understood by health facility staff. If donors flood the local health system with additional staff, withdrawing them will result in less sustainable outcomes (26).

## Limitations

Our results should be considered, in addition to their limitations. First, our data are from government health facilities and NGOs operating in South Africa’ s Western Cape, which had the lowest estimated prevalence of HIV among 15-49-year-olds (9.4%; 95% CI: 8.5, 10.2%) in 2013, compared to the other provinces, such as KwaZulu-Natal (26.8%; 95% CI: 25.8%–27.6%).(29) Therefore, our results may not be generalizable to other provinces and/or other sub-Saharan African countries where PEPFAR was implemented. Second, our data was aggregate data that provided by Tier.net, a government-owned HIV electronic patient management system. As such, we run the risk of possible loss of information and are vulnerable to the ecological fallacy, resulting in false inferences about individual behavior on the basis of population-level data.

However, our estimates of retention are consistent with previously published literature on retention in ART programs in South Africa’ s Western Cape during PEPFAR (74.2%; 95% CI: 73.2%–75.2%) (30) and post-PEFAR 54.3% (95% CI:52.4%-56.1%) at 36 months follow up that used individual patient level data in their analysis (31). Third, we could have unmeasured confounding due to the inability to control for potential confounders due to missing information at the facility level (i.e., transfers, employee turnover rate). We attempted to minimize confounding by type of health facility by including only primary healthcare facilities (i.e., clinics, community day centers, and community health centers) in our study.

## Conclusion

Our results show that when donor funding decreased, there was a decline in RIC of patients in HIV care post-PEPFAR compared to the PEPFAR direct service era. To ensure that the RIC is high, the system needs to minimize loss to follow-up. Support from different government bodies and the size of health facilities had no effect on RIC, although additional human resources in the system and support from NGOs with a history in the province assisted in sustaining retention. Although it is unlikely that there would ever be as large a program as PEPFAR to support HIV care and treatment in the future, it would be valuable for donors working in government health facilities to allocate funding to support health facilities and NGOs as they transition out. These funds would help maintain quality patient care and sustain clinical outcomes during the transition. In conclusion, future donor transitions should prioritize close planning with local governments, together with stable human and financial resources, to ensure sustained health outcomes.

## Data Availability

All data produced in the present work are contained in the manuscript

