## Supplemental Figure 2 for "Evaluation of the impact of PEPFAR transition on retention in care in South Africa’s Western Cape"

**Supplemental Figure 2a-2e. 12 month retention in care, loss to follow-up and death stratified by NGO and overlaid on total patients on ART at the start and end of the 12-month period.**

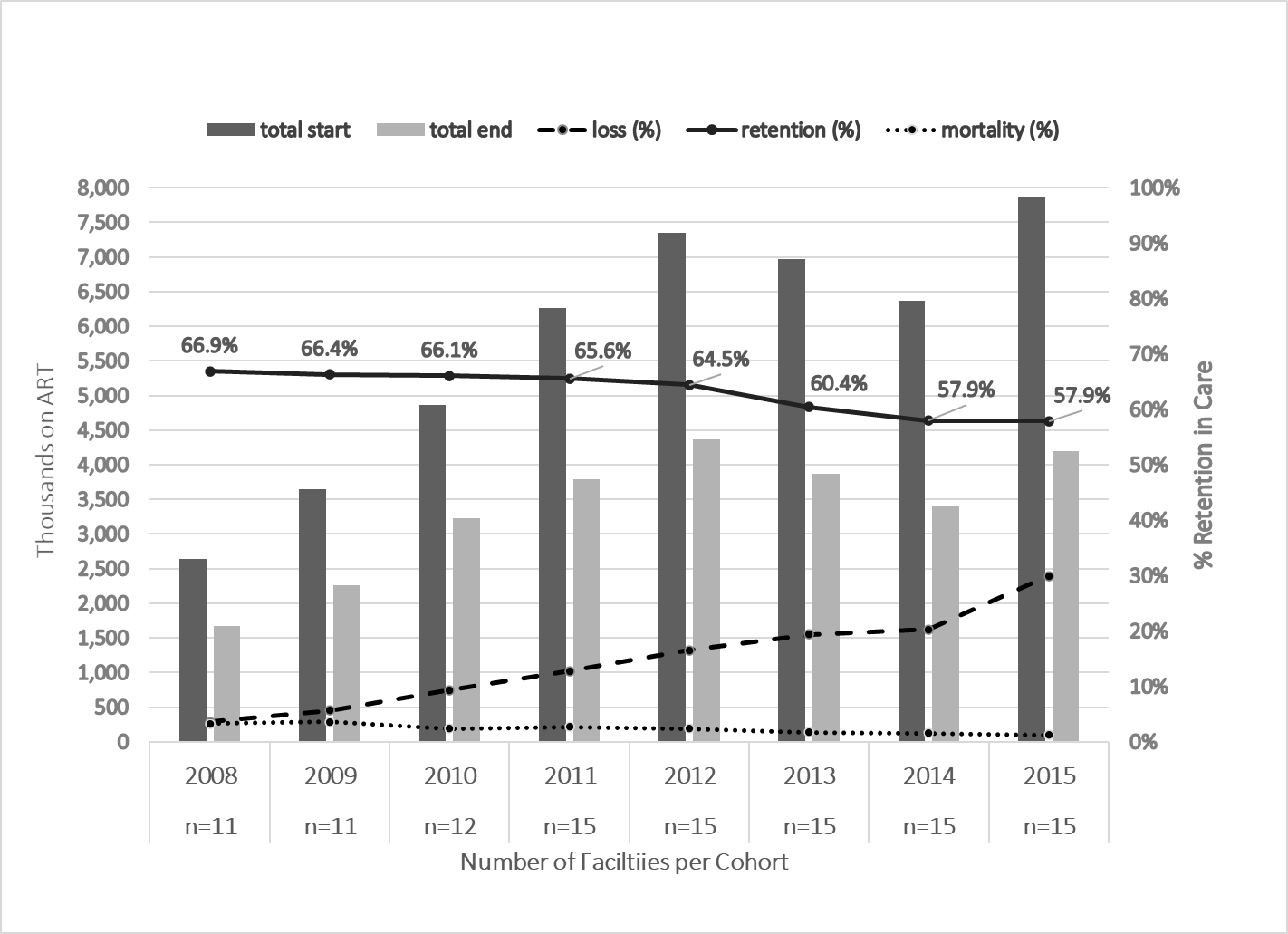

a) **Anova/Kheth’Impilo**

**d) Anova**

b) **that’sIt**

**b) Kheth’Impilo**

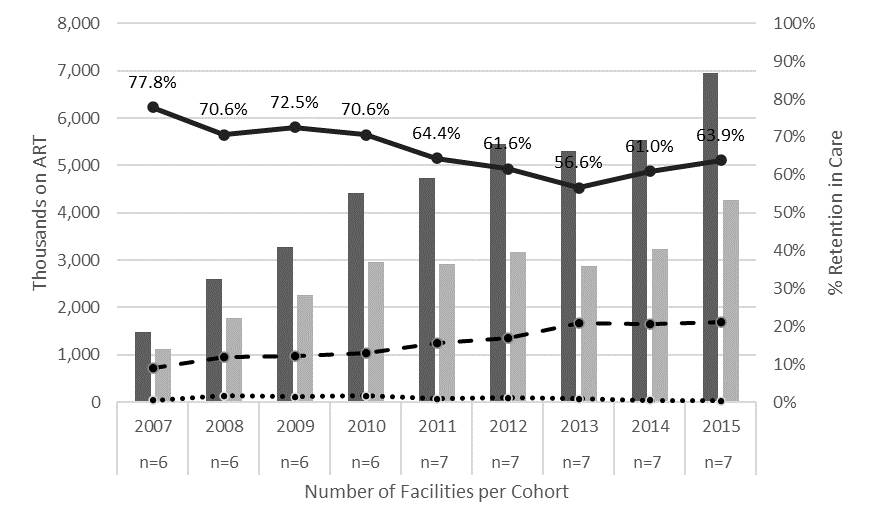

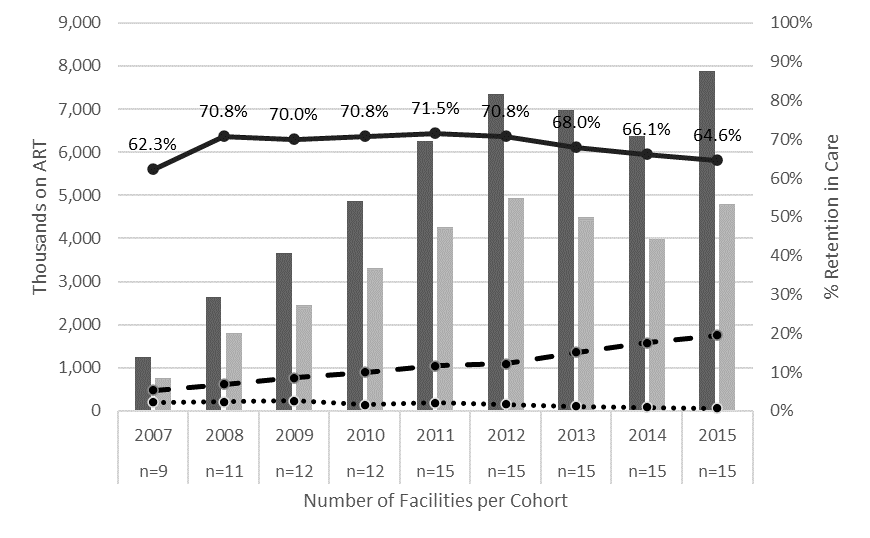

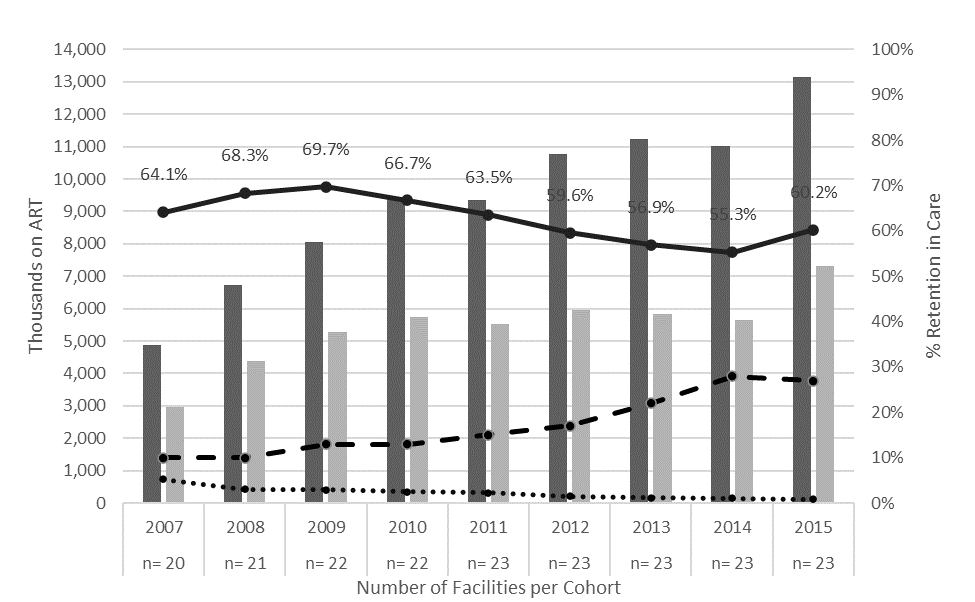

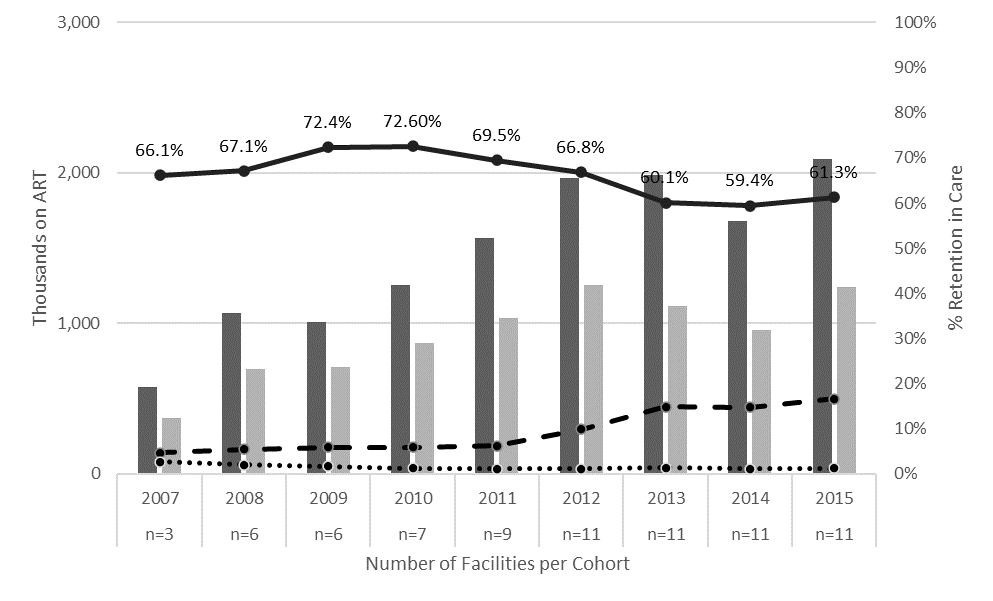

**E	e) Right to Care**

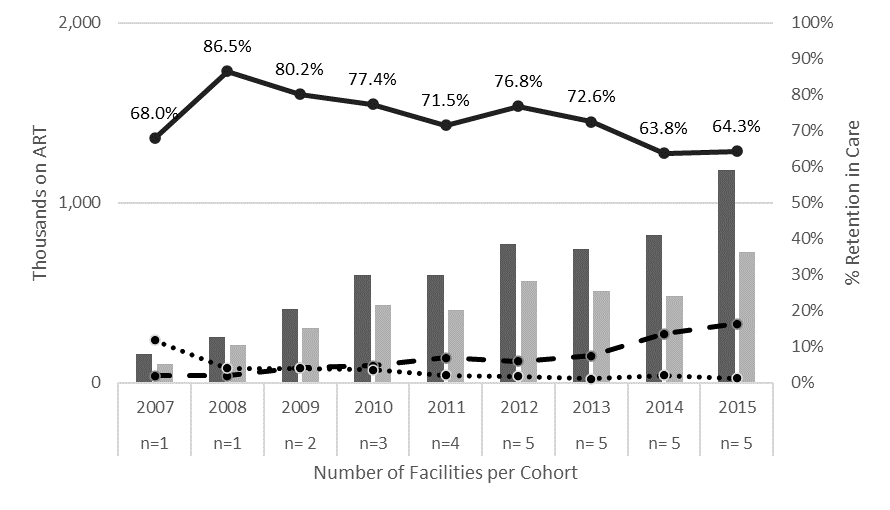
