## Supplemental Table 1 for "Evaluation of the impact of PEPFAR transition on retention in care in South Africa’s Western Cape"

**Supplement Table 1. Crude and adjusted quantile regression for the outcome of 24-month retention.**

|  | **Crude quantile difference (95% CI)** | | | **Adjusted quantile difference (95% CI)** | | |
| --- | --- | --- | --- | --- | --- | --- |
|  | **25%** | **50%** | **75%** | **25%** | **50%** | **75%** |
| **post PEPFAR** | -5.3% (-10.3, -0.2%) | -5.8% (-9.7, -2.0%) | -3.5% (-7.7, 0.6%) | -5.8% (-11.5, -0.2%) | -6.9% (-10.2, -3.7%) | -1.6% (-6.2, 3.1%) |
| **slope during PEPFAR** | -1.0% (-2.0, -0.0%) | -1.2% (-2.0, -0.4%) | -2.0% (-2.8, -1.2%) | -0.5% (-1.7, 0.6%) | -1.2% (-1.9, -0.6%) | -2.8% (-3.7, -1.9%) |
| **change in slope post-PEPFAR** | 0.8% (-3.1, 4.7%) | 0.5% (-2.5, 3.5%) | 0.4% (-2.8, 3.5%) | 0.5% (-3.9, 4.9%) | 0.9% (-1.6, 3.4%) | 0.7% (-3.0, 4.3%) |
| **constant** | 54.5% (50.9, 58.2%) | 60.4% (57.6, 63.2%) | 63.5% (60.5, 66.5%) | 47.9% (40.1, 55.7%) | 65.5% (61.1, 70.0%) | 67.8% (61.4, 74.3%) |
| **PEPFAR NGO** |  |  |  |  |  |  |
| Anova (constant) | - | - | - | ref. | ref. | ref. |
| Kheth'impilo | - | - | - | 10.0% (4.7, 15.2%) | 1.1% (-1.8, 4.1%) | 1.1% (-3.2, 5.5%) |
| Anova/ Kheth'impilo | - | - | - | 3.6% (-2.2, 9.4%) | -2.8% (-6.2, 0.5%) | -2.8% (-7.6, 2.0%) |
| Right to Care | - | - | - | 9.2% (1.7, 16.8%) | 8.1% (3.8, 12.4%) | 9.4% (3.1, 15.6%) |
| that'sit | - | - | - | 5.5% (-0.1, 11.0%) | 6.0% (2.8, 9.2%) | 7.6% (3.0, 12.2%) |
| **clinic size** |  |  |  |  |  |  |
| 174-1020 (constant) | - | - | - | ref. | ref. | ref. |
| 1021-2562 | - | - | - | 7.7% (2.1, 13.3%) | 2.5% (-0.7, 5.7%) | 3.2% (-1.5, 7.8%) |
| 2563-4021 | - | - | - | 7.4% (1.7, 13.1%) | 1.5% (-1.8, 4.7%) | -0.9% (-5.7, 3.8%) |
| 4022-5856 | - | - | - | 7.2% (1.4, 13.1%) | 0.9% (-2.5, 4.2%) | 0.8% (-4.1, 5.6%) |
| 5857-9760 | - | - | - | 10.0% (3.1, 16.9%) | 0.8% (-3.1, 4.8%) | -2.8% (-8.6, 2.9%) |
| **Location** |  |  |  |  |  |  |
| urban (constant) | - | - | - | ref. | ref. | ref. |
| rural | - | - | - | 4.9% (-1.4, 11.2%) | -2.5% (-6.0, 1.1%) | -5.0% (-10.2, 0.3%) |
| **government support** |  |  |  |  |  |  |
| City of Cape Town (constant) | - | - | - | ref. | ref. | ref. |
| Western Cape Government | - | - | - | -7.2% (-12.0, -2.6%) | -7.9% (-10.6, -5.2%) | -8.2% (-12.1, -4.3%) |
| Combined | - | - | - | 2.5% (-6.3, 11.3%) | -5.0% (-10.0, 0.1%) | -6.5% (-13.8, 0.8%) |
