## Supplemental Table 2 for "Evaluation of the impact of PEPFAR transition on retention in care in South Africa’s Western Cape"

**Supplemental Table 2. Basic quantile regression models for the outcome of 12- and 24-month retention**

|  | **Basic quantile difference (95% CI) 12-month** | | | | | | | **Basic quantile difference (95% CI) 24-month** | | | | | | |
| --- | --- | --- | --- | --- | --- | --- | --- | --- | --- | --- | --- | --- | --- | --- |
|  | **25%** | **25%** | | **25%** | | **25%** | | | **25%** | | **25%** | **25%** | | **25%** |
| **12 month retention** |  |  | |  | |  | | |  | |  |  | |  |
| **post PEPFAR** | -6.9% (13.5, -0.2%) | -5.9% (-11.4, -0.4%) | | -6.0% (-12.6, 0.5%) | | -4.5% (-11.3, 2.4%) | | | -4.5% (-10.1, 1.1%) | | -5.2% (-9.6, -0.7%) | -5.4% (-10.5, -0.4%) | | -6.7% (-11.6, -1.8%) |
| **slope during PEPFAR** | 0.4% (-1.0, 1.7%) | 0.5% (-0.6, 1.6%) | | 0.3% (-1.0, 1.6%) | | 0.4% (-0.9, 1.8%) | | | -0.5% (-1.6, 0.6%) | | -0.6% (-1.5, 0.3%) | -0.7% (-1.7, 0.3%) | | -0.4% (-1.4, 0.5%) |
| **change in slope post-PEPFAR** | 0.6% (-4.6, 5.8%) | 0.1% (-4.1, 4.4%) | | -0.4% (-5.5, 4.6%) | | -0.9% (-6.2, 4.4%) | | | 0.1% (-4.3, 4.4%) | | 0.2% (-3.3, 3.7%) | 0.1% (-3.8, 4.1%) | | 0.5% (-3.3, 4.3%) |
| **constant** | 58.4% (53.0, 63.8%) | 56.7% (52.0, 61.3%) | | 65.0% (60.0, 7.0%) | | 71.1% (65.0, 77.3%) | | | 52.7% (48.2, 57.2%) | | 51.6% (47.7, 55.4%) | 57.3% (53.4, 61.3%) | | 63.4% (59.0, 67.8%) |
| **PEPFAR NGO** |  |  | |  | |  | | |  | |  |  | |  |
| Anova | ref. | - | | - | | - | | | ref. | | - | - | | - |
| Kheth'impilo | 8.7% (4.2, 13.3%) | - | | - | | - | | | 7.8% (4.0, 11.6%) | | - | - | | - |
| Anova/ Kheth'impilo | 5.3% (-0.5, 11.1%) | - | | - | | - | | | 2.1% (-2.8, 6.9%) | | - | - | | - |
| that'sit | 3.9% (-3.7, 11.5%) | - | | - | | - | | | 3.7% (-2.6, 10.1%) | | - | - | | - |
| Right to Care | -0.8% (-6.1, 4.6%) | - | | - | | - | | | 0.8% (-3.6, 5.3%) | | - | - | | - |
| **Health Facility size** |  |  | |  | |  | | |  | |  |  | |  |
| 174-1020 | - | ref. | | - | | - | | | - | | ref. | - | | - |
| 1021-2562 | - | 4.5% (-0.1, 9.1%) | | - | | - | | | - | | 3.4% (-0.3, 7.2%) | - | | - |
| 2563-4021 | - | 1.6% (-3.1, 6.3%) | | - | | - | | | - | | 1.8% (-2.0, 5.6%) | - | | - |
| 4022-5856 | - | 12.3% (7.6, 16.9%) | | - | | - | | | - | | 8.6% (4.8, 12.4%) | - | | - |
| 5857-9760 | - | 6.0% (1.2, 10.7%) | | - | | - | | | - | | 3.6% (-0.3, 7.4%) | - | | - |
| **Location** |  |  | |  | |  | | |  | |  |  | |  |
| Urban | - | - | | ref. | | - | | | - | | - | ref. | | - |
| Rural | - | - | | -6.0% (-9.5, -2.5%) | | - | | | - | | - | -3.7% (-6.4, -1.0%) | | - |
| **government support** |  |  | |  | |  | | |  | |  |  | |  |
| City of Cape Town | - | - | | - | | ref. | | |  | | - | - | | ref. |
| Western Cape Gov | - | - | | - | | -1.5% (-11.8, 8.8%) | | |  | | - | - | | -8.5% (-11.8, -5.2%) |
| Combined | - | - | | - | | -11.3% (-15.9, 6.7%) | | |  | | - | - | | -2.8% (-10.2, 4.6%) |
|  | **50%** | **50%** | | **50%** | | **50%** | | | **50%** | | **50%** | **50%** | | **50%** |
| **12 month retention** |  |  | |  | |  | | |  | |  |  | |  |
| **post PEPFAR** | -4.5% (-9.4, 0.4%) | -5.4% (-9.8, -1.0%) | | -5.1% (-9.7, -0.6%) | | -5.3% (-9.1, -1.4%) | | | -5.0% (-8.4, -1.7%) | | -5.6% (-9.3, -1.9%) | -5.5% (-9.3, -1.7%) | | -5.8% (-9.3, 2.3%) |
| **slope during PEPFAR** | -1.4% (-2.3, -0.4%) | -0.8% (-1.7, 0.1%) | | -1.0% (-1.9, -0.0%) | | -1.1% (-1.9, -0.3%) | | | -1.7% (-2.3, -1.0%) | | -1.1% (-1.9, -0.4%) | -1.2% (-1.9, -0.4%) | | -1.3% (-2.0, -0.6%) |
| **change in slope post-PEPFAR** | 0.5% (-3.3, 4.3%) | 0.1% (0.0, 3.4%) | | 0.2% (-3.3, 3.8%) | | 0.2% (-3.3., 3.8%) | | | 1.1% (-1.5, 3.7%) | | 0.8% (-2.1, 3.7%) | 0.1% (-2.8, 3.1%) | | 0.3% (-2.4, 3.0%) |
| **constant** | 63.6% (59.7, 67.6%) | 63.7% (60.0, 67.4%) | | 70.0% (66.0, 73.1%) | | 74.8% (71.3, 78.3%) | | | 55.8% (53.1, 58.5%) | | 59.2% (56.0, 62.3%) | 61.9% (59.0, 64.9%) | | 66.0% (62.8, 69.2%) |
| **PEPFAR NGO** |  | |  | |  | |  |  | |  | | |  | |
| Anova | ref. | - | | - | | - | | | ref. | | - | - | | - |
| Kheth'impilo | 5.9% (2.6, 9.2%) | - | | - | | - | | | 6.4% (4.1, 8.6%) | | - | - | | - |
| Anova/ Kheth'impilo | 0.8% (-3.4, 5.1%) | - | | - | | - | | | 2.7% (-0.3, 5.6%) | | - | - | | - |
| that'sit | 0.6% (-5.0, 6.2%) | - | | - | | - | | | 6.6% (2.8, 10.4%) | | - | - | | - |
| Right to Care | 1.8% (-2.1, 5.8%) | - | | - | | - | | | 3.8% (1.1, 6.4%) | | - | - | | - |
| **clinic size** |  |  | |  | |  | | |  | |  |  | |  |
| 174-1020 | - | ref. | | - | | - | | | - | | ref. | - | | - |
| 1021-2562 | - | 1.5% (-2.2, 5.2%) | | - | | - | | | - | | -0.3% (-3.4, 2.8%) | - | | - |
| 2563-4021 | - | 1.2% (-2.5, 5.0%) | | - | | - | | | - | | 0.5% (-2.7, 3.6%) | - | | - |
| 4022-5856 | - | 8.8% (5.1, 12.5%) | | - | | - | | | - | | 4.4% (-1.3, 7.6%) | - | | - |
| 5857-9760 | - | 3.6% (-0.2, 7.4%) | | - | | - | | | - | | 0.1% (-3.1, 3.3%) | - | | - |
| **Location** |  |  | |  | |  | | |  | |  |  | |  |
| Urban | - | - | | ref. | | - | | | - | | - | ref. | | - |
| Rural | - | - | | -5.5% (-8.0, -3.1%) | | - | | | - | | - | -3.3% (-5.3, -1.3%) | | - |
| **government support** |  |  | |  | |  | | |  | |  |  | |  |
| City of Cape Town | - | - | | - | | ref. | | | - | | - | - | | ref. |
| Western Cape Gov | - | - | | - | | -3.7% (-9.5, 2.1%) | | | - | | - | - | | -7.6% (-9.9, -5.2%) |
| Combined | - | - | | - | | -10.1% (-12.7, -7.6%) | | | - | | - | - | | -5.2% (-10.5, 0.1%) |
|  | **75%** | **75%** | | **75%** | | **75%** | | | **75%** | | **75%** | **75%** | | **75%** |
| **12 month retention** |  |  | |  | |  | | |  | |  |  | |  |
| **post PEPFAR** | -5.2% (-8.7, -1.6%) | -4.5% (-8.3, -0.7%) | | -5.3% (-9.1, -1.5%) | | -6.4% (-10.5, -2.4%) | | | -5.2% (-9.4, -1.0%) | | -4.5% (-8.9, 0.0%) | -5.2% (-9.2, -1.2%) | | -5.0% (-8.5, -1.5%) |
| **slope during PEPFAR** | -1.8% (-2.6, -1.2%) | -1.6% (-2.4, -0.9%) | | -1.5% (-2.3, -0.8%) | | -1.5% (-2.3, -0.7%) | | | -2.3% (-3.2, -1.5%) | | -2.1% (-3.0, -1.2%) | -2.0% (-2.8, -1.2%) | | -2.1% (-2.8, -1.4%) |
| **change in slope post-PEPFAR** | -3.6% (-2.7, 2.9%) | 0.0% (-3.0, 3.0%) | | -0.6% (-3.6, 2.3%) | | 1.9% (-0.4, 4.2%) | | | 1.0% (-2.3, 4.3%) | | 0.8% (-2.7, 4.3%) | 0.4% (-2.7, 3.5%) | | 0.0% (-2.7, 2.7%) |
| **constant** | 69.5% (66.7, 72.4%) | 73.4% (70.1, 76.6%) | | 74.9% (72.0, 77.9%) | | 77.9% (74.2, 81.6%) | | | 60.4% (57.0, 63.8%) | | 64.9% (61.1, 68.7%) | 65.6% (62.5, 68.7%) | | 69.2% (66.1, 72.4%) |
| **PEPFAR NGO** |  |  | |  | |  | | |  | |  |  | |  |
| Anova | ref. | - | | - | | - | | | ref. | | - | - | | - |
| Kheth'impilo | 5.2% (2.8, 7.6%) | - | | - | | - | | | 6.2% (3.3, 9.1%) | | - | - | | - |
| Anova/ Kheth'impilo | 0.2% (-2.9, 3.3%) | - | | - | | - | | | 2.1% (-1.5, 5.8%) | | - | - | | - |
| that'sit | 2.7% (-1.3, 6.7%) | - | | - | | - | | | 6.1% (1.3, 10.9%) | | - | - | | - |
| Right to Care | 0.4% (-2.4, 3.2%) | - | | - | | - | | | 3.5% (0.1, 6.8%) | | - | - | | - |
| **Health facility size** |  |  | |  | |  | | |  | |  |  | |  |
| 174-1020 | - | 73.4% (70.1, 76.6%) | | - | | - | | | - | | 64.9% (61.1, 68.7%) | - | | - |
| 1021-2562 | - | -4.5% (-7.7, -1.3%) | | - | | - | | | - | | -1.9% (-5.6, 1.9%) | - | | - |
| 2563-4021 | - | -3.1% (-6.4, 0.1%) | | - | | - | | | - | | -3.0% (-6.9, 0.8%) | - | | - |
| 4022-5856 | - | 1.2% (-2.1, 4.4%) | | - | | - | | | - | | 1.4% (-2.4, 5.2%) | - | | - |
| 5857-9760 | - | -1.9% (-5.2, 1.4%) | | - | | - | | | - | | -2.5% (-6.3, 1.4%) | - | | - |
| **Location** |  |  | |  | |  | | |  | |  |  | |  |
| Urban | - | - | | 74.9% (72.0, 77.9%) | | - | | | - | | - | 65.6% (62.5, 68.7%) | | - |
| Rural | - | - | | -5.3% (-7.3, -3.2%) | | - | | | - | | - | -3.1% (-5.2, -0.9%) | | - |
| **government support** |  |  | |  | |  | | |  | |  |  | |  |
| City of Cape Town | - | - | | - | | 77.9% (74.2, 81.6%) | | | - | | - | - | | 69.2% (66.1, 72.4%) |
| Western Cape Gov | - | - | | - | | -4.2% (-10.3, 1.9%) | | | - | | - | - | | -7.2% (-9.5, -4.8%) |
| Combined | - | - | | - | | -8.1% (-10.8, -5.3%) | | | - | | - | - | | -6.2, -11.5, -1.0%) |
