## Supplementary figures and images for "Evaluation of the impact of PEPFAR transition on retention in care in South Africa’s Western Cape"

### Supplemental Figure 1

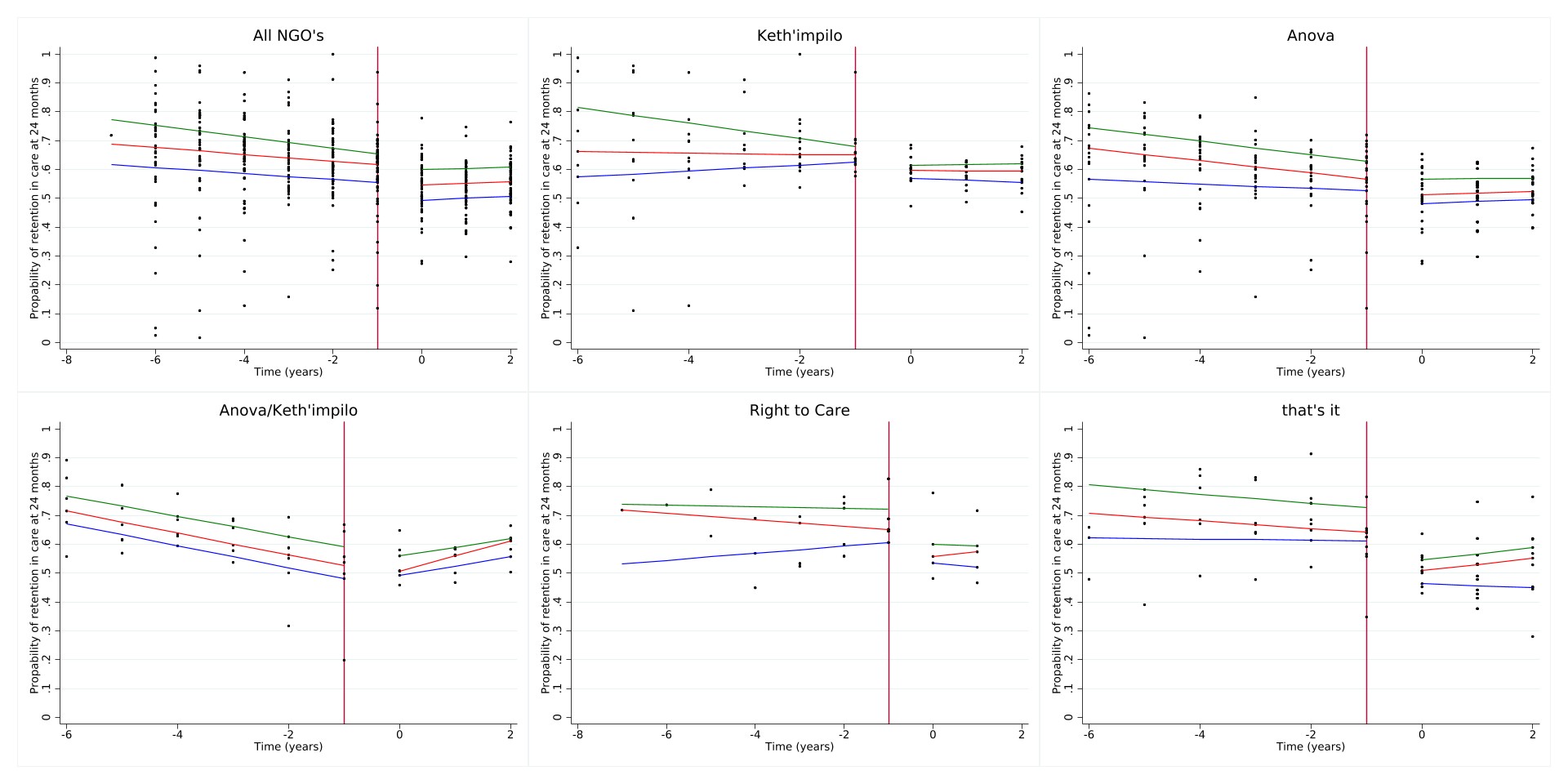
**Supplemental Figure 1a-1f. Quantile regression estimates for 24-month retention stratified by NGO**
